# A Tale of Two Populations: A meta-analysis of hemodynamic, procedural, and clinical outcomes of PASCAL Ace for Mitral and Tricuspid Regurgitation

**DOI:** 10.1101/2025.06.05.25329087

**Authors:** Sebastian Balda, Ivo Diaz-Djevoich, Matias Panchana-Lascano, Flavio Veintemilla-Burgos, Geovanna Minchalo-Ochoa, Thomas Leone, Mohammed Al-Tawil

## Abstract

**Background:** Mitral and tricuspid regurgitation (MR, TR) cause significant morbidity and mortality, especially in high-surgical-risk patients. The PASCAL Ace transcatheter edge-to-edge repair (TEER) device shows promise for treating both, yet existing evidence is limited by single-center studies and heterogeneous cohorts. This single-arm meta-analysis systematically evaluates the safety, efficacy, and clinical impact of PASCAL Ace.

**Methods:** A comprehensive literature search (PubMed, Scopus, Embase) identified studies reporting hemodynamic, procedural, and clinical outcomes of PASCAL Ace for MR/TR. Observational cohorts and randomized trials were included; systematic reviews, case reports, studies with other devices, and non-English publications were excluded. Data extracted included demographics, baseline parameters, procedural details, and outcomes. Study quality was assessed (Newcastle-Ottawa Scale). Meta-analysis used a random-effects model to pool proportions and weighted mean differences, with heterogeneity (I²) evaluated. Significance was p<0.05 (Stata 18).

**Results:** Seven studies (324 patients: 195 MR, 129 TR) were included. Baseline symptom burden was high (nearly all NYHA III/IV: 100% TR, 94.9% MR). Pooled overall mortality was 4% (95% CI 1%–6%, I²=0.01%). Procedural success was 96% (95% CI 94%–99%, I²=12.53%). Post-procedure, 84% achieved NYHA I/II status (95% CI 77%–90%, I²=0.00%), alongside significant reductions in regurgitation severity. Major adverse events occurred in 4% (95% CI 1%–6%, I²=0.01%).

**Conclusions:** The PASCAL Ace device effectively improves functional status (evidenced by NYHA class reduction) in patients with MR and TR, though outcomes were comparatively more favorable in MR. The persistence of right heart dysfunction in TR underscores the need for studies on complementary therapies and long-term follow-up to optimize results. This meta-analysis supports PASCAL Ace as a safe and efficacious TEER option.

## Introduction

Mitral and tricuspid regurgitation (MR, TR) are prevalent valvular heart diseases associated with significant morbidity and mortality. (1,2) The development of transcatheter edge-to-edge repair (TEER) has provided alternative therapeutic options for patients who have considered high risk for surgical intervention. (3) The PASCAL Ace system, a novel TEER device, has been developed to address the limitations of earlier technologies and enhance procedural outcomes. (4) The PASCAL Ace is a refined version of the original PASCAL system. It approximates valve leaflets using two clasps and paddles surrounding a central spacer, which fills the regurgitant orifice to decrease the regurgitation. The system allows for independent or simultaneous leaflet capture, and its ability to elongate facilitates safe retrieval from the left ventricle, minimizing the risk of chordal entanglement. (5)

The design makes PASCAL Ace suitable for anatomically challenging cases, such as small mitral valve areas (MVAs), commissural jets, large flail gaps (>10 mm), and redundant leaflet tissue, as seen in Barlow’s disease. (6) Moreover, it features Narrower Paddles of 6 mm compared to 10 mm in the original; a Smaller Central Spacer, which allows a greater leaflet approximation relative to implant size; Increased Paddle Curvature, which enhances the neo coadaptation area and distributes tension across the captured leaflet tissue. (5) PASCAL Ace is effective in treating both degenerative and functional MR, especially in patients with complex anatomies or smaller MVAs. Its design allows for precise leaflet capture and tension distribution, leading to significant MR reduction. (7) In an early compassionate use study, PASCAL Ace achieved MR grade ≤2 in 93% of patients in one year, with 88% survival and 94% freedom from heart failure hospitalization. (8)

Moreover, it has been utilized for tricuspid TEER. (9) The PASCAL Ace system is particularly suited for patients with complex tricuspid anatomies, including those with large coaptation gaps or challenging leaflet orientations. Its design allows for effective treatment in cases where traditional surgical approaches may be high-risk or contraindicated. (10) A meta-analysis involving 549 patients reported an 83.5% procedural success rate, with significant improvements in NYHA functional class, six-minute walk distance, and TR severity over follow-up periods ranging from 30 days to one year. (9)

These findings highlight the potential of the PASCAL Ace system as a valuable transcatheter treatment for both MR and TR, demonstrating favorable procedural success rates and significant clinical improvements. However, most existing studies have been limited to single-center experiences or heterogeneous cohorts, making it essential to consolidate evidence through a meta-analytic approach. This study aims to conduct a single-arm meta-analysis to systematically evaluate the hemodynamic, procedural, and clinical outcomes of the PASCAL Ace system in patients with MR and TR. By synthesizing available data, we seek to provide a comprehensive assessment of its safety and efficacy, aiding clinical decision-making and future research directions.

## Materials and Methods

### Literature Search

A comprehensive and detailed literature search was conducted across well-known and widely used databases, including PubMed, Scopus, and Embase. The most relevant studies meeting our inclusion and exclusion criteria were selected. The search term used was “PASCAL ACE”. The protocol for this meta-analysis was registered in PROSPERO (International Prospero Register of Systematic Reviews) under the ID CRD420251009196.

### Inclusion and Exclusion Criteria

The Rayyan platform was used for screening and selecting eligible studies. Three co-authors independently conducted the initial screening of study titles and abstracts, with any selection conflict resolved by an additional reviewer.

Inclusion criteria for the study included observational cohort studies and randomized controlled trials that compared hemodynamic, procedural and clinical outcomes between two populations: mitral and tricuspid regurgitation.

The exclusion criteria contained systematic review studies, meta-analyses, narrative reviews, case reports/series, editorials, study protocols, commentaries, letters, and non-English studies were excluded. In addition, patients younger than 18 years old were excluded. Finally, patients without tricuspid or mitral valve regurgitation or who were treated with any other device that isn’t PASCAL Ace, were also excluded.

### Data extraction

A data extraction sheet was elaborated using Google Sheets by one co-author. One co-author extracted data independently for every 2 articles. The data collected involved first author last name, the year of publication, location where the study was conducted, the study design, the number of patients, and demographic characteristics, such as sex, age, Body Mass Index (BMI), European System for Cardiac Operative Risk Evaluation (EuroSCORE) II, Pre-procedure NYHA score, baseline Tricuspid Annular Plane Systolic excursion (TAPSE), baseline RV diameter (mm), severity of regurgitation grade, previous coronary artery disease, presence of atrial fibrillation, implanted ICD, diabetes mellitus, hypertension, Chronic Obstructive Pulmonary Disease, renal failure, previous stroke and previous cardiac surgery.

The collected outcomes included all time mortality rate, device success, post procedure NYHA score, post procedure valve regurgitation grade, major complications. Continuous outcomes collected included the number of implanted clips, post procedure systolic Pulmonary Artery pressure, post procedure mean transvalvular pressure gradient, hospital length stay, RV diameter (mm), TAPSE (mm), procedure time (min).

The quality of the studies was assessed by using the New-Castle Ottawa Scale, following established guidelines, and it can be visualized in the Supplementary Digital Content, as Table S1.

### Statistical Analysis

The pooling of proportions was carried out with a random effects model. For continuous outcomes we used an inverse variance approach to determine the weighted mean difference. Heterogeneity was evaluated using I2 statistics, with I2 > 50% indicating substantial heterogeneity. A p value of less than 0.05 was statistically significant. The statistical analysis was done using Stata 18 (2023).

## Results

### Study characteristics

Seven studies were included for the final analysis. Three studies included patients treated with PASCAL Ace for tricuspid regurgitation, three for mitral regurgitation, and one study included both types of valves. Search strategy is demonstrated in Figure 1. A total of 324 patients were included, from which 129 were intervened for tricuspid, and 195 for mitral. In the tricuspid group, 42.3% of the patients were female, with a mean age of 76 ± 4.4. In the mitral group, 42.7% were female, and the patients had a mean age of 79 ± 2.21. All the patients with tricuspid regurgitation presented a NYHA Score III or IV before the procedure, very similar to 94.9% of the patients with mitral regurgitation. The rest of baseline characteristics included studies and demographic data from the patients, are shown in Tables 1, 2 and 3.

**Figure 1.**
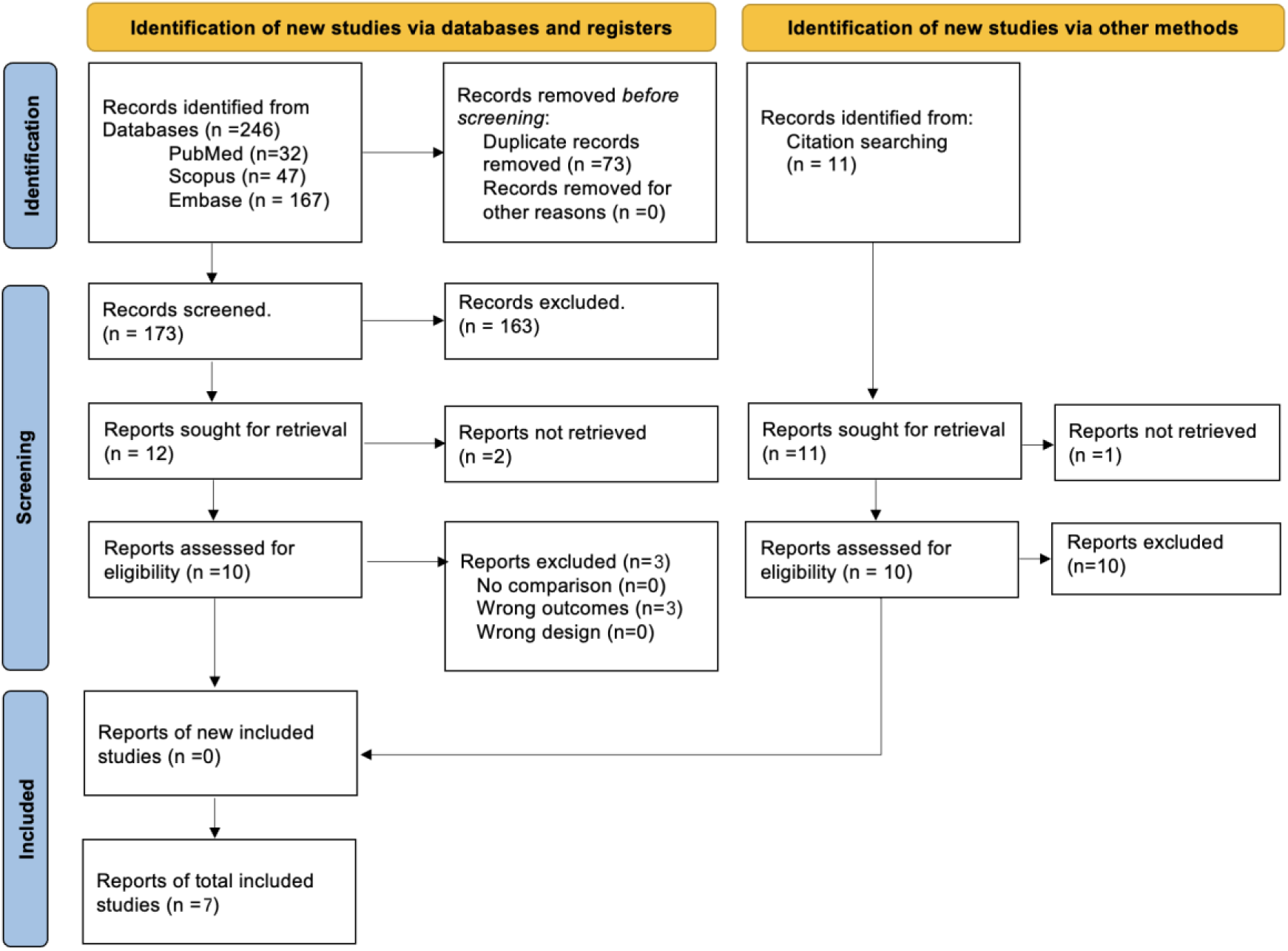
PRISMA flowchart that shows search strategy used for this study

**Table 1.** Characteristics of included studies.

| Study ID | Location | Duration | Design | No. of patients |  | Study conclusions |
| --- | --- | --- | --- | --- | --- | --- |
|  |  |  |  | Tricuspid | Mitral |  |
| **Aurich et al, 2021** | University Hospital of Heidelberg. Heidelberg, Germany | 2020 | Retrospective Cohort | 16 |  | The system offers an interventional treatment option that expands the spectrum of devices for percutaneous tricuspid valve leaflet repair. |
| **Rottlander et al, 2022** | Multicenter. Cologne, Germany | 2020-2021 | Retrospective Cohort | 20 | 20 | PASCAL Ace demonstrated excellent reliability compared to the standard monitoring using a dedicated catheter with sideholes |
| **Volz et al, 2022** | University Hospital of Heidelberg. Heidelberg, Germany | 2020-2021 | Retrospective Cohort | 11 |  | Interventional direct leaflet repair in TR with the PASCAL Ace device improve the VO2 max |
| **Wild et al, 2022** | Multicenter. Germany and Switzerland | 2017-2021 | Retrospective Cohort | 82 |  | The PASCAL Ace repair system has high technical and procedural success rates with efficient TR reduction and significant clinical and echocardiographic improvement at follow-up. |
| **Barth et al, 2022** | Cardiovascular Center Bad Neustadt. Saxony-Anhalt, Germany | 2012-2016 | Retrospective Cohort |  | 66 | PASCAL Ace is safe and effective in treating MR, resulting in a sustained MR reduction, a reverse cardiac remodelling, improvement of exercise capacity, quality of life, NT-proBNP levels and hemodynamics at follow-up. |
| **Moonen et al, 2022** | Multicenter. Australia and Canada | 2017-2023 | Retrospective Cohort |  | 17 | At 1 year, the PASCAL Ace implant system demonstrated feasibility in this early, compassionate use experience in a small group of symptomatic patients with anatomically complex MR |
| **Paukovitch et al, 2024** | University Heart Center Ulm. Ulm, Germany | 2017-2019 | Retrospective Cohort |  | 72 | The use of the PASCAL Ace device in patients with small MVOAs might correlate with a risk of both clinically relevant orifice reduction and even iatrogenic stenosis. |

**Table 2.** Baseline characteristics of patients included in the studies; EuroSCORE II, European System for Cardiac Operative Risk Evaluation; NYHA,New York Heart Association Classification.

| Study ID | Valve | Female (%) | Age (Mean ± SD) | EuroSCORE II (Mean ± SD) | Pre procedure NYHA score |  |  |
| --- | --- | --- | --- | --- | --- | --- | --- |
|  |  |  |  |  | I-II | III | IV |
| Aurich et al, 2021 | Tricuspid | 4 (25) | 73 ± 11 | 7.2 ± 4 | 0 | 12 | 4 |
| Rottlander et al, 2022 | Tricuspid and Mitral | 13 (65) | 79.9 ± 1.7 | 7.4 ± 1.2 | 0 | 17 | 3 |
| Volz et al, 2022 | Tricuspid | 3 (27.3) | 71 ± 9 | 5.5 ± 3.7 | 0 | 10 | 1 |
| Wild et al, 2022 | Tricuspid | 36 (43.9) | 79 ± 6 | 7.6 ± 6 | 0 | 72 | 10 |
| Barth et al, 2022 | Mitral | 24 (36.4) | 80 ± 18 | 29.8 ± 16 | 0 | 54 | 5 |
| Moonen et al, 2022 | Mitral | 6 (35.3) | 76 ± 13 | 5.7 ± 4.7 | 7 | 10 | 0 |
| Paukovitsch et al, 2024 | Mitral | 18 (65) | 81 ± 2.73 | 6.2 ± 1.85 | 0 | 25 | 15 |

**Table 3.** Baseline characteristics of patients included in the studies; COPD,chronic obstructive pulmonary disease.

| Study ID | Atrial fibrillation (%) | COPD (%) | Renal failure (%) | Coronary artery disease (%) | Pre procedure regurgitation (%) |  | Permanent pacemaker implanted (%) |
| --- | --- | --- | --- | --- | --- | --- | --- |
|  |  |  |  |  | Grade 3 | Grade 4 |  |
| Aurich et al, 2021 | 10 (62.5) | 4 (25) | 7 (73.8) | 9 (56.3) | 3 (18.8) | 13 (81.3) | 7 (43.8) |
| Rottlander et al, 2022 | 20 (100) | 1 (5) | 1 (5) | 7 (35) | 7 (35) | 13 (65) | - |
| Rottlander et al, 2022 | 20 (100) | 1 (5) | - | 7 (35) | 7 (35) | 13 (65) | - |
| **Volz et al, 2022** | 7 (63.6) | 4 (36.4) | 5 (45.5) | 8 (72.7) | 3 (27.3) | 8 (72.7) | 5 (45.5) |
| **Wild et al, 2022** | - | - | 14 (17.1) | - | - | 37 (45.1) | - |
| **Barth et al, 2022** | 43 (65.2) | - | - | - | 28 (42.4) | 38 (57.6) | 3 (4.5) |
| **Moonen et al, 2022** | 11 (64.8) | 5 (29.4) | 8 (47.1) | 8 (47.1) | 5 (29.4) | 12 (70.6) | 6 (35.3) |
| **Paukovitsch et al, 2024** | 26 (65) | 4 (10) | 30 (75) | 29 (72.5) | 7 (67.5) | 13 (32.5) | 0 |

### Procedural Outcomes

Our results show the device success rate with the PASCAL Ace system was 98% in patients with MR (95% CI: 95–100%, I² = 0.00%) and 85% in those with TR (95% CI: 67–104%, I² = 92.81%) illustrated in Figure 2. Overall, the total device success rate was 96% (95% CI: 94– 99%, I² = 12.53%). Furthermore, the pooled prevalence of all-cause mortality was 4% (95% CI: 1–6%, I² = 0.01%), with no significant differences observed between valve groups, as shown in Figure 3. It is important to note that Aurich et al, Wild et al, and Barth et al reported 30-day mortality, Rottlandeer et al reported a 3-month mortality, and finally Moonen et al a 1-year mortality. Moreover, MR patients required fewer implanted clips on average (1.28 ± 0.45) compared to TR patients (1.79 ± 0.56). On the other hand, major complications were rare and similar between groups, occurring in 2.1% of MR cases and 2.3% of TR cases. Lastly, Procedure time, weighted by the number of patients, was shorter in the MR cohort (69.6 ± 40.2 minutes) compared to the TR cohort (98.6 ± 53.6 minutes) as shown in Table S2.

**Figure 2.**
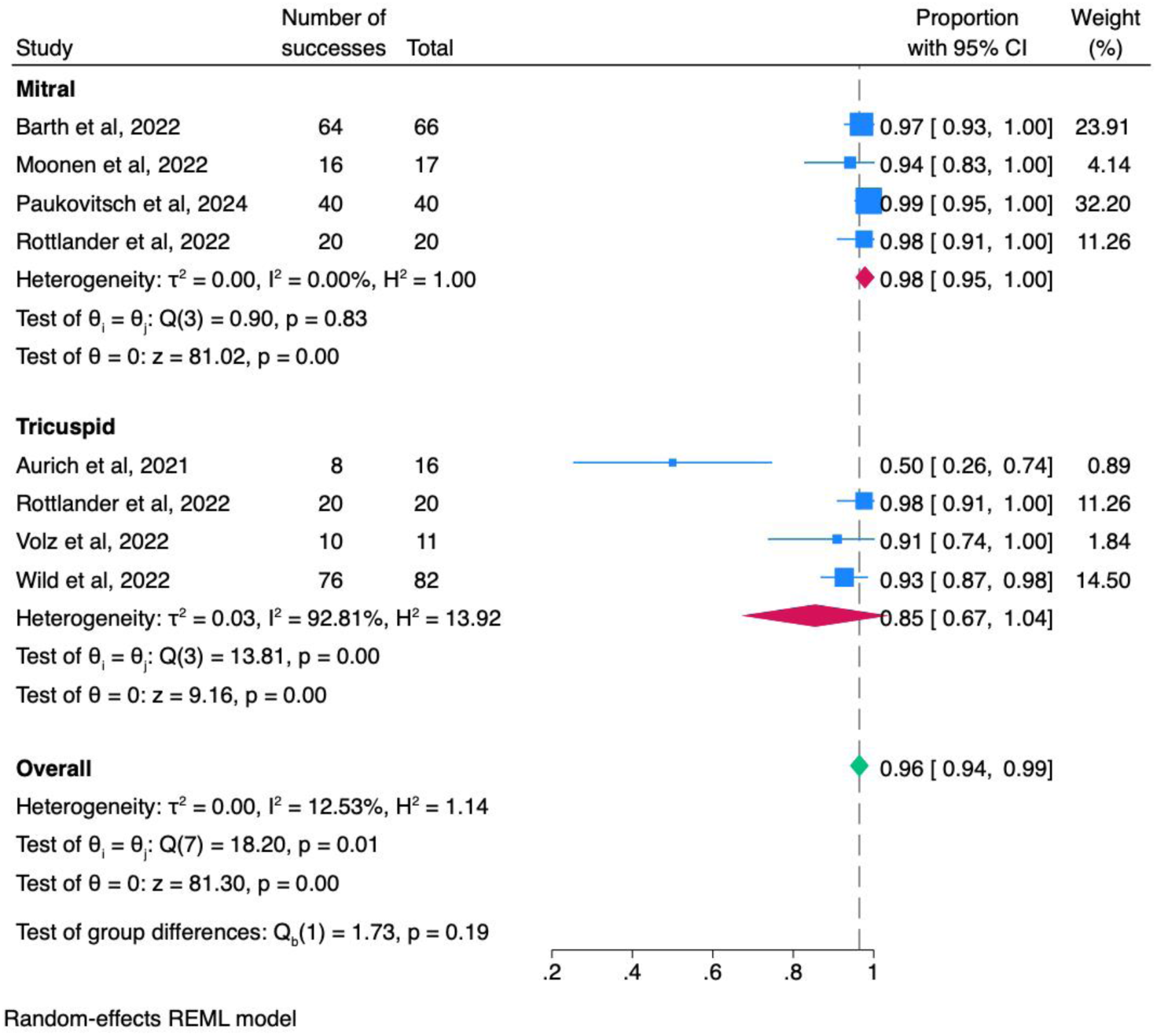
Forest plot of pooled prevalence of device success rates among included studies

**Figure 3.**
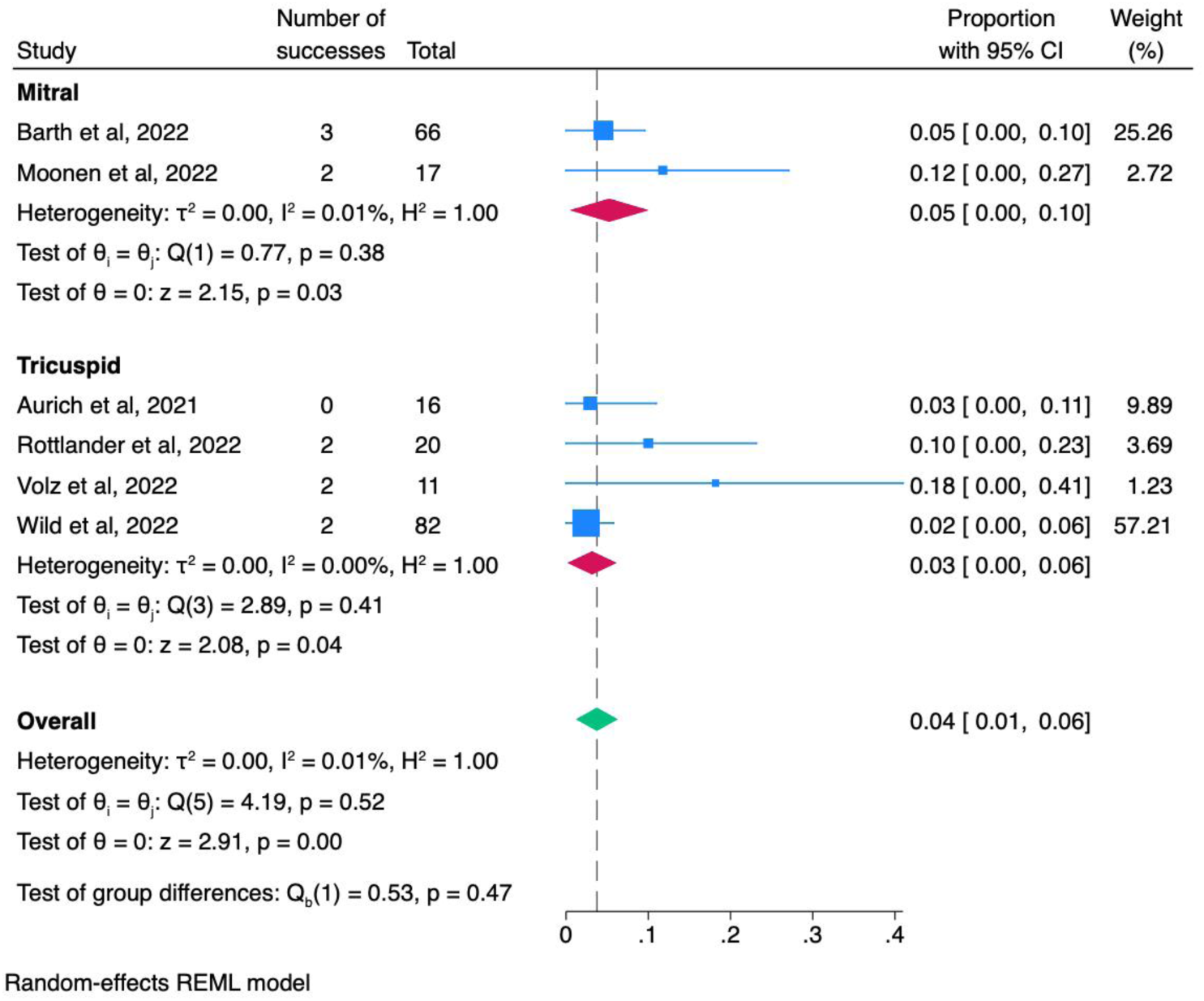
Forest plot of pooled prevalence of mortality rates among included studies

### Clinical Outcomes

#### NYHA

Regarding improvement in NYHA functional classification following TEER with the PASCAL Ace system, findings are shown in Figure 4 and Table S3. At baseline, the vast majority of patients were categorized as NYHA Class III or IV, indicating significant symptomatic limitation. Post-procedure follow-up revealed a marked shift in functional status, with 84% (95% CI: 77–90%, I² = 0.00%) of patients achieving NYHA Class I or II, reflecting substantial symptom relief and improved exercise tolerance. This improvement was observed across both MR and TR cohorts, although MR patients demonstrated a greater degree of functional recovery, with 87% (95% CI: 79–94%, I² = 0.01%) reaching Class I–II, compared to 71% (95% CI: 51–91%, I² = 50.56%) in the TR group. Notably, no patients remained in NYHA Class IV at follow-up, and only a minority persisted in Class III (13% MR vs. 29% TR), underscoring the clinical efficacy of the intervention. These findings support the role of PASCAL Ace in significantly enhancing quality of life for patients with advanced valvular diseases.

**Figure 4.**
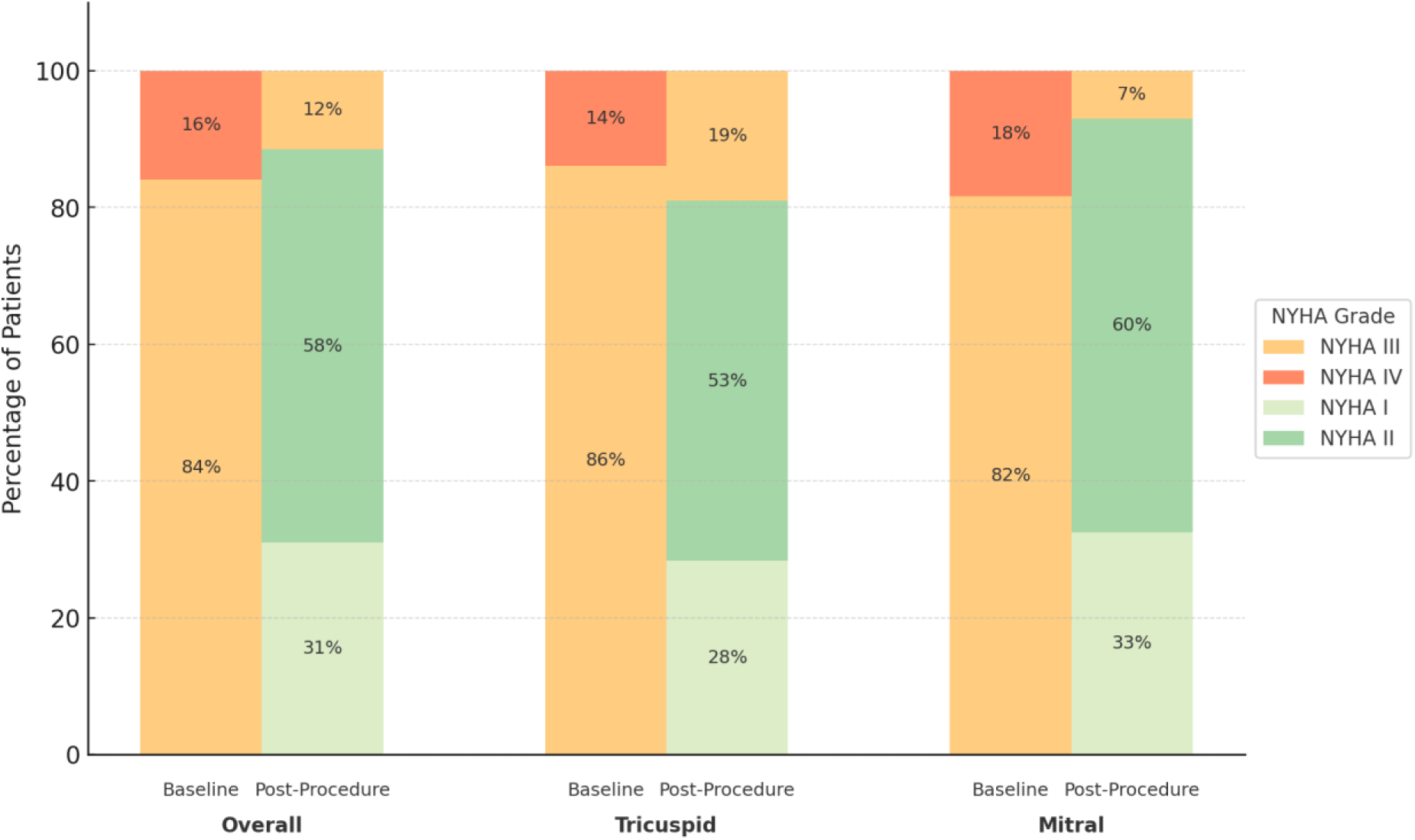
NYHA grade at baseline and Post-procedure follow up; NYHA,New York Heart Association classification.

#### Valve Regurgitation Severity

About the severity of mitral and tricuspid valve regurgitation, pre- and post-procedure parameters show improvement, as shown in Figure 5 and Table S4. Initially, nearly all patients presented with Grade III or IV regurgitation. After the procedure, 62% (95% CI: 47– 78%, I² = 85.54%) of patients achieved minimal or no residual regurgitation (Grade 0–I), while 33% (95% CI: 18–47%, I² = 84.29%) improved to Grade II. Only a small minority remained at Grade III, and no patients were classified as Grade IV post-intervention. These findings reflect the effectiveness of the device in both mitral and tricuspid regurgitation, with consistent outcomes across both populations. The pronounced shift toward lower grades of regurgitation underscores the role of PASCAL Ace in achieving meaningful anatomical correction and symptomatic benefit.

**Figure 5.**
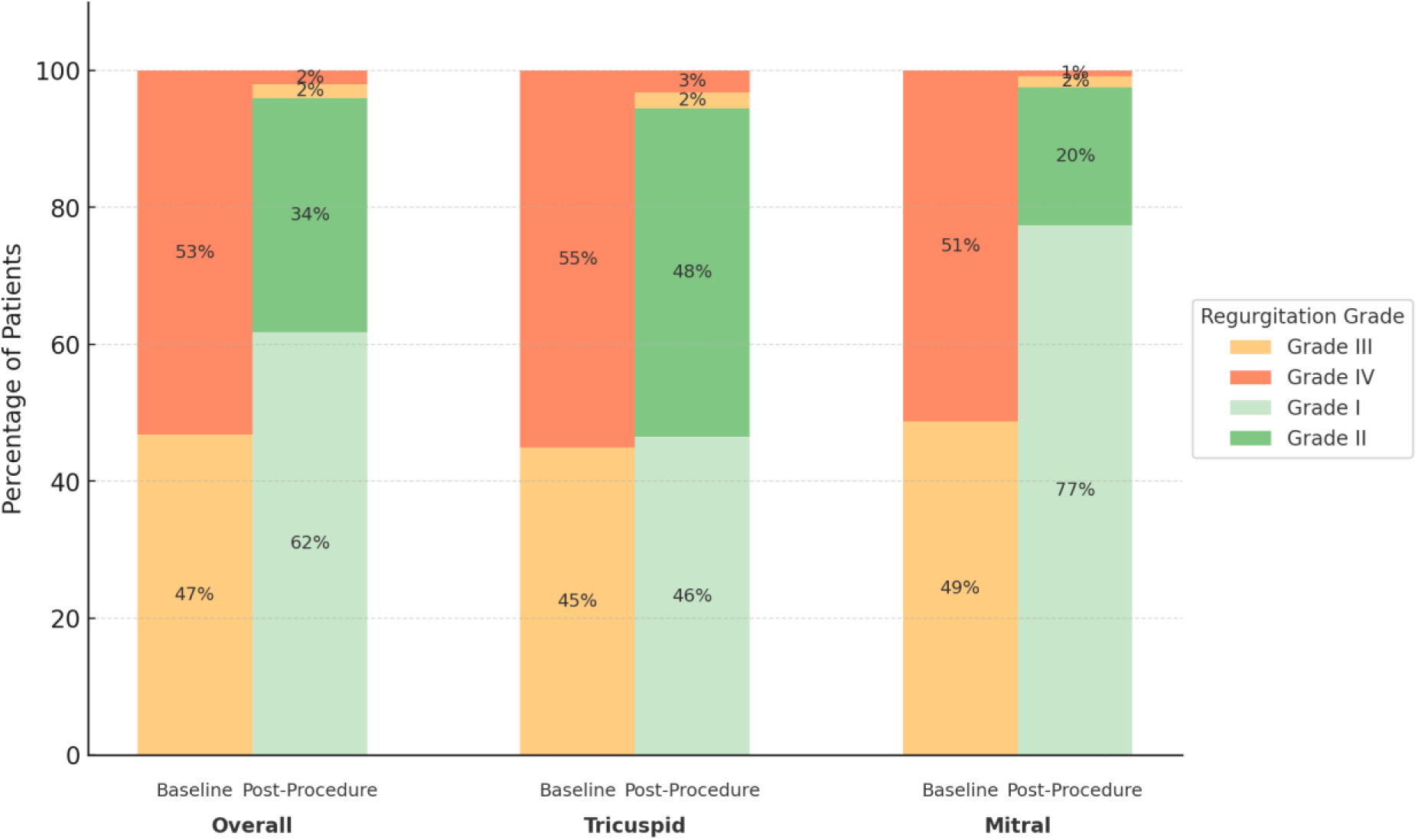
Severity of regurgitation grade at baseline and Post-procedure follow up.

PASCAL Ace demonstrated high efficacy in reducing regurgitation severity in both mitral and tricuspid populations. In patients with MR, the majority initially presented with Grade III or IV disease. Following the procedure, 67% of MR patients achieved Grade 0–I regurgitation, and 31% reached Grade II, while none remained in Grade IV and only 2% in Grade III. This resolution highlights the device’s effectiveness in promoting leaflet coaptation and performance within the mitral anatomy.

A meaningful reduction was similarly observed among patients with TR. At baseline, most of the patients were in Grade III or IV; post-procedure, 55% improved to Grade 0–I and 38% to Grade II, with only 7% remaining in Grade III and none in Grade IV. Although the degree of regurgitation reduction was slightly less pronounced than in MR, these outcomes remain clinically significant. Especially taking into consideration the anatomical complexity of the tricuspid valve and the frequent presence of right heart dysfunction. Collectively, these findings reinforce the efficacy of PASCAL Ace in achieving significant anatomical and hemodynamic improvements across both valvular pathologies.

#### Hemodynamic outcomes

For the analysis of hemodynamic outcomes, we included studies with complete pre and post procedure data shown in Table 4 which yielded the following results; first, post RV diameter mild reduction from a baseline of 45.3 ± 7.2 mm to a post procedure 43.1± 6.5 mm. Second, TAPSE stability from 16.3± 2.4 mm at baseline to 16.8 ± 1.9 mm post procedure. Third, the systolic Pulmonary artery pressure did not change significantly from baseline 47.1±7.2 mmHg to post procedure 46.6 ±8.1mmHg.

**Table 4.** Comparison of hemodynamic outcomes of patients with tricuspid regurgitation, pre and post procedure, treated with PASCAL Ace; RV, Right Ventricle; TAPSE, Tricuspid Annular Plane Systolic Excursion; PA, Pulmonary Artery.

| Study ID | Baseline RV Diameter | Post procedure RV Diameter | Baseline TAPSE | Post Procedure TAPSE | Baseline Systolic PA Pressure | Post procedure Systolic PA Pressure |
| --- | --- | --- | --- | --- | --- | --- |
| **Aurich et al, 2021** | 50 ± 6 | 50 ± 7 | 13± 1.7 | 15 ± 2.5 | - | - |
| **Rottlander et al, 2022** | 37±1.9 | 36 ± 1.7 | 19±1.5 | 18 ± 0.9 | 46 ± 3.3 | 47 ± 2.8 |
| **Volz et al, 2022** | 49± 6 | 45 ± 7 | 16± 0.4 | 15 ± 0.4 | 49 ± 11 | 46 ± 13 |
| **Wild et al, 2022** | 55 ± 11 | - | 18 ± 4 | - | 42 ± 19 | - |

## Discussion

The findings of this study provide insights regarding the safety and efficacy of the PASCAL Ace system for the treatment of mitral and tricuspid regurgitation. For instance, the high procedural success rates and significant improvements in clinical and hemodynamic outcomes across both patient populations. Our results are in accordance with previous studies, further strengthening the potential use of PASCAL Ace as a viable alternative to surgical interventions.

Our study reports samples with either left or right heart disease, being tricuspid and mitral regurgitation respectively. The complete set of patients had severe functional impairment evidenced by a NYHA III or IV in all the TR patients and most of the MR patients (94.9%). However, when reviewing the surgical outcomes separately, it can be said that procedures on MR patients had fewer chip implantations and had less surgical time than their TR counterparts. The device success was high among both populations, with MR at the lead with 98 % followed by TR with 85%. This 13% difference, fewer clips, and shorter procedures may be due to the complexity of tricuspid valve anatomy and the functional associated right heart failure, which complicates the interventions in tricuspid regurgitation. During the follow-up period, patients with TR showed no improvement on the diameter of the right ventricle, TAPSE, and maintained elevated systolic pressure in the pulmonary artery. These findings support the theory of a persistent right heart failure in this population of patients. This could be explained as a chronic volume overload from severe TR, taking time to reverse (11). Hence, the benefits of reduced regurgitation may become more apparent with longer follow-up.

MR patients treated with PASCAL Ace showed more improvement regarding severity of the regurgitation than the TR counterpart. A possible explanation could be that Mitral regurgitation can be structurally repaired while TR is frequently functional and associated with right ventricle dysfunction and remodeling (12)(13).

Our Study found excellent results among both groups for improving the regurgitation grade with PASCAL Ace, slightly favoring the mitral population. Reducing regurgitation from grade III to II can decrease RV volume overload and improve RV function over time. As demonstrated in our study, 50% of patients achieved Grade I regurgitation, which is associated with minimal symptoms and near-normal cardiac function, improving quality of life and reducing pulmonary hypertension.

Patients with MR showed greater improvement in NYHA functional class than those with TR. 87% of patients with MR achieved class I-II, compared with 71% of patients with TR. This difference may be attributed to persistent right ventricular dysfunction in TR, which is not fully addressed by valve repair. Despite these improvements, 29% of patients with TR and 13% of patients with MI remained in class III, reflecting persistent cardiac dysfunction and the need for adjuvant therapies.

A key strength of our analysis is the systematic evaluation of both MR and TR populations, highlighting the versatility of the PASCAL Ace system. The pooled procedural success rate of 96% underscores the reliability of this device in achieving successful valve implantation. Moreover, the improvement in NYHA functional class and reduction in regurgitation severity post-procedure reflect the significant symptomatic relief provided by PASCAL Ace. These improvements are critical for enhancing patients’ quality of life and reducing the burden of disease.

Despite these promising results, several limitations must be acknowledged. First, the heterogeneity among included studies, particularly regarding follow-up durations and outcome assessments, poses a challenge in drawing uniform conclusions. While the overall mortality rate was low (4%), differences in reporting periods, ranging from 30 days to one-year, complicate direct comparisons. Additionally, the reliance on observational studies introduces inherent biases, and the lack of randomized controlled trials limits the robustness of our findings.

Furthermore, variations in patient selection criteria and operator experience across studies may have influenced procedural outcomes. Future large-scale, multicenter randomized trials are warranted to confirm these findings and establish standardized protocols for the use of PASCAL Ace in clinical practice.

## Conclusions

The PASCAL Ace system provides a high procedural success rate and significant improvements in clinical outcomes for patients with MR and TR. Our meta-analysis supports its role as an effective transcatheter therapy, particularly in high-risk surgical candidates. However, further prospective randomized studies are needed to validate these findings and refine patient selection criteria to optimize clinical outcomes. The consolidation of real-world data and long-term follow-up studies will be instrumental in guiding future therapeutic strategies and expanding the applicability of PASCAL Ace in valvular heart disease management.

## Authors’ Consent for Publication

We, the authors of the manuscripts entitled “A Tale of Two Populations:A meta-analysis of hemodynamic, procedural, and clinical outcomes of PASCAL Ace for Mitral and Tricuspid Regurgitation”, hereby declare that we have participated in the conception, design, and execution of the research work. We confirm that we have reviewed the final version of the manuscript and approved it for submission to *Circulation: Cardiovascular Interventions*. Furthermore, we understand that this submission implies that the manuscript, in whole or in part, has not been published previously and is not under consideration for publication elsewhere.

We affirm that all listed authors have agreed to be named as contributors to this work and have read and approved the final manuscript.

Sincerely,

Sebastian BALDA MD,

Ivo DIAZ-DJEVOICH MD,

Matias PANCHANA-LASCANO MD,

Flavio VEINTEMILLA-BURGOS MD,

Geovanna MINCHALO-OCHOA MD,

Thomas LEONE MD,

Mohammed AL-TAWIL MD

## Data Availability

The datasets and materials used during this research are available upon reasonable request. Interested researchers can contact Mohammed Al-Tawil, MD for access to the data and materials supporting the findings of this study.

## Acknowledgements

We would like to express our heartfelt appreciation to everyone who contributed to the completion of this research. Their valuable support, guidance, and encouragement have been instrumental in shaping this work.

Lastly, our deep appreciation goes to our families and friends for their understanding, encouragement, and unwavering belief in our abilities throughout this journey.

Thank you for your support.

**Permission to reproduce material from other sources:** Not applicable.

## Clinical trial registration

Not applicable.

## Ethics Approval

Not applicable

## Authorship confirmation/contribution statement

**Sebastian BALDA:**

Conceptualization, Methodology, Validation, Formal analysis, Supervision, Visualization, Project administration, Funding acquisition, Writing - original draft, Writing - review & editing, Investigation, Data curation

**Thomas LEONE:**

Conceptualization, Methodology, Validation, Formal analysis, Supervision, Project administration, Writing - review & editing

**Flavio VEINTEMILLA-BURGOS:**

Conceptualization, Methodology, Validation, Formal analysis, Supervision, Visualization, Project administration, Funding acquisition, Writing - original draft, Writing - review & editing, Investigation, Data curation

**Mathias PANCHANA-LASCANO:**

Conceptualization, Methodology, Data curation, Investigation, Validation, Visualization, Writing - original draft, Writing - review & editing, Formal analysis, Project administration, Supervision

**Rodolfo KRONFLE:**

Conceptualization, Methodology, Validation, Investigation, Formal analysis, Writing - review & editing, Writing - original draft, Visualization, Data curation

**Geovanna MINCHALO-OCHOA:**

Conceptualization, Methodology, Validation, Investigation, Formal analysis, Writing - review & editing, Writing - original draft, Visualization, Data curation

**Ivo DIAZ-DJEVOICH:**

Conceptualization, Methodology, Validation, Investigation, Formal analysis, Writing - review & editing, Writing - original draft, Visualization, Data curation

**Mohammed AL-TAWIL:**

Conceptualization, Methodology, Validation, Investigation, Formal analysis, Writing - review & editing, Writing - original draft, Visualization, Data curation, Supervision

## Supplementary

**Table S1.** Risk of Bias Assessment using the NewCastle-Ottawa Scale.

| Study ID | Selection |  |  |  | Comparability |  | Outcome |  |  | Score |
| --- | --- | --- | --- | --- | --- | --- | --- | --- | --- | --- |
|  | Representativ-<br>ness of the<br>exposed | Selection<br>of Non-Exposed | Ascertainm-<br>ent of<br>Exposure | Outcome of<br>Interest Not<br>Present at Start<br>of Study | Main<br>Factor | Additional<br>Factor | Assess-<br>ment | Follow-up<br>Lenght | Adequa-<br>cy of<br>follow-up |  |
| Aurich et al, 2021 | * | 0 | * | * | * | * | * | * | 0 | 7 |
| Rottlander et al, 2022 | * | 0 | * | * | * | * | * | * | 0 | 8 |
| Volz et al, 2022 | * | 0 | * | * | * | * | * | * | * | 8 |
| Wild et al, 2022 | * | 0 | * | * | * | * | * | * | * | 8 |
| Barth et al, 2022 | * | 0 | * | * | * | * | * | * | * | 8 |
| Moonen et al, 2022 | * | 0 | * | * | * | * | * | * | * | 8 |
| Paukovitsch et al, 2024 | * | 0 | * | * | * | * | * | 0 | 0 | 6 |

**Table S2.**
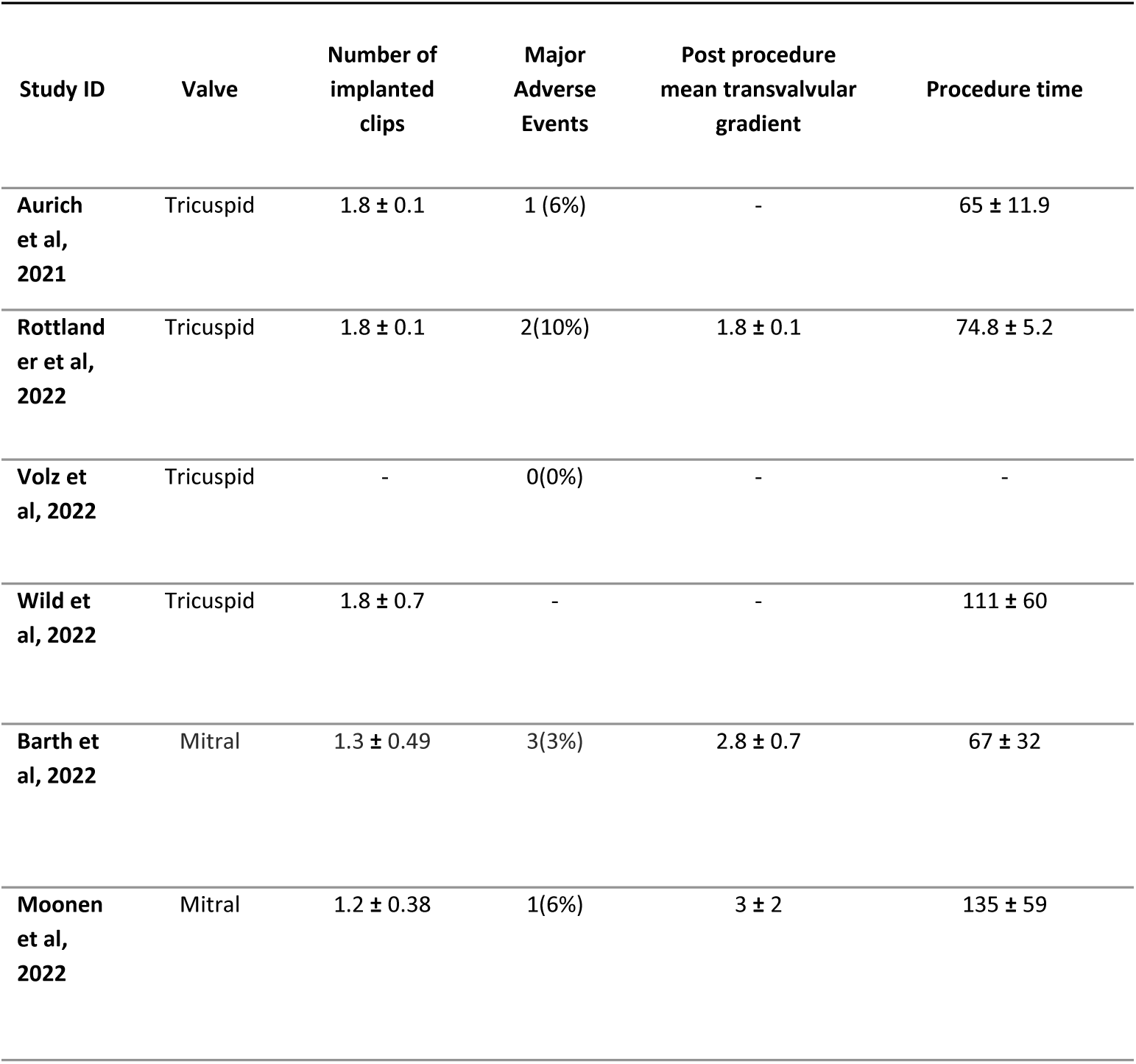
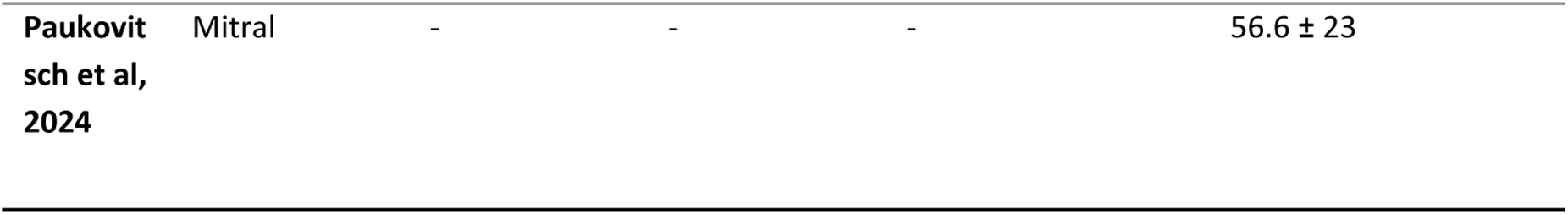
Secondary postoperative outcomes of patients treated with PASCAL Ace.

**Table S3.** NYHA baseline and outcomes of patients treated with PASCAL Ace; NYHA, New York Heart Association classification.

| Study ID | Valve | Baseline NYHA Grade III | Baseline NYHA Grade IV | Post Procedure NYHA Grade III | Post Procedure NYHA Grade I-II | NYHA Grade improvement from Baseline (%) |
| --- | --- | --- | --- | --- | --- | --- |
| **Aurich et al, 2021** | Tricuspid | 12 | 4 | 1 | 10 | 69 |
| **Rottlander et al, 2022** | Tricuspid | 17 | 3 | 3 | 17 | 85 |
| **Volz et al, 2022** | Tricuspid | 10 | 1 | 4 | 7 | 73 |
| **Wild et al, 2022** | Tricuspid | 72 | 10 | - | - | - |
| **Barth et al, 2022** | Mitral | 54 | 5 | 3 | 51 | 87 |
| **Moonen et al, 2022** | Mitral | 10 | 0 | 2 | 15 | 88 |
| **Paukovitsch et al, 2024** | Mitral | 25 | 15 | - | - | - |

**Table S4.** Comparison of regurgitation grades pre and post procedure, with patients treated with Pascal ACE.

| Study ID | Valve | Baseline Severity of Regurgitation Grade III | Baseline Severity of Regurgitation Grade ≥IV | Post Procedure Severity of Regurgitation Grade ≤I | Post Procedure Severity of Regurgitation Grade II | Post Procedure Severity of Regurgitation Grade III | Post Procedure Severity of Regurgitation Grade IV | Severity of regurgitation Grade Improvement from Baseline (%) |
| --- | --- | --- | --- | --- | --- | --- | --- | --- |
| Aurich et al, 2021 | Tricuspid | 3 | 13 | 8 | 2 | 1 | 3 | 77 |
| Rottlander et al, 2022 | Tricuspid | 7 | 13 | 8 | 11 | 1 | 0 | 95 |
| Volz et al, 2022 | Tricuspid | 3 | 8 | 6 | 3 | 1 | 1 | 82 |
| Wild et al, 2022 | Tricuspid | 45 | 37 | 37 | 45 | 0 | 0 | 100 |
| Barth et al, 2022 | Mitral | 28 | 38 | 53 | 12 | 1 | 0 | 100 |
| Moonen et al, 2022 | Mitral | 5 | 12 | 8 | 8 | 1 | 1 | 89 |
| Paulkovitsch et al, 2024 | Mitral | 27 | 13 | 35 | 5 | 0 | 0 | 100 |

